# REPAIR OF ABDOMINAL WALL DEFECTS WITH ACELLULAR BOVINE PERICARDIUM MATRIX A MULTICENTRE PROSPECTIVE STUDY

**DOI:** 10.1101/2021.06.15.21257792

**Authors:** Luiz Fernando Frascino, Jonas Dias De Campos Severi, Fernanda Ribeiro Funes Lorenzzato, Hamilton Luiz Xavier Funes

## Abstract

**Background:** The association of prosthetic meshes in the abdominal wall repair, reducing the recurrence rates in an impactful way, has become an almost mandatory routine for the success of these surgeries. After decades using non-biological synthetic implants, from the 90s onwards biological membranes of animal or human origin were introduced – the so called acellular biological matrices - beginning a new era in abdominal wall defects correction.

**Methods:** Thirty patients underwent repair for different abdominal wall deformities, with acellular matrices of bovine pericardium, in a total of 40 anatomically individualized implants. The median follow-up was 22 months, with patients evaluated clinically and radiologically. In three cases, biopsies of the implanted areas were performed, allowing histological analysis of the material.

**Results:** There was no recurrence of hernias in any of the cases, both clinically and radiologically. There was also no record of hematomas, infections or any phenomenon of a local or systemic reaction nature. Radiologically, it was not possible to visualize the matrices at the implantation site in any of the analysed postoperative periods. Biopsies showed important tissue neoformation replacing the implanted membranes, with important deposition of collagen, normal-looking cellularized tissue, and absence of foreign body reactions.

**Conclusions:** The analysed matrices showed similarity to other biological membranes described in the international literature. Representing an important update and conceptual evolution, biological matrices must be incorporated into the therapeutic arsenal in abdominal wall repairs.

## Introduction

The impairment of anatomical and functional integrity of the muscular abdominal wall is a relatively frequent occurrence, with varying degrees of clinical manifestation, complexity and causal agent, mainly represented by hernias in their different forms, laxity or exaggerated bulging of the anterior abdominal plane and sequelae after trauma, infections or tumour resections.

Public data from Brazil show an annual average of 242.000 herniorrhaphies, with 54% corresponding to inguinal hernias, with 99.4% being by open approach and only 0.6% by laparoscopy, with 22% reported as operated in an emergency situation^1^. Added to this, there are patients operated on private services and repairs secondary to other causes, indicating the magnitude of the problem and requiring an algorithm for the appropriate selection of the technique to be adopted in the different corrections^2^.

In addition, diffuse bulging due to loosening of the abdominal wall muscles, with functional and aesthetic impairment, is a common and not properly estimated occurrence in post-bariatric patients, in repetitive pregnancies or in android biotype women, affecting quality of life and work capacities in a much larger portion of the population, with great demand for surgical treatment. Although complete in its integrity, these repairs generally require reinforcement with prosthetic meshes after muscle correction due to the thinning of the structures, thus preventing the protrusion recurrence, the main factor to negatively impact long-term outcomes^3^.

Although comorbidity factors such as obesity, diabetes and smoking among others can affect the recurrence rates^4^, the association of biomaterials allowed to repair different abdominal wall defects with important tension relief, concomitant to increasing local resistance, reducing the recurrence rates by more than 50%, evidencing the mechanical factor as preponderant^5^ and making the use of reinforcement meshes in these situations a mandatory routine.

Thus, after decades using non-biological synthetic meshes^6^, from the 90s onwards, biological scaffolds from animal or human origin were introduced - called acellular biological matrices - thus beginning a new era in the repair of abdominal wall defects^7^, with results stimulating its growing adoption around the world in the past two decades.

Different acellularized membranes have been developed from varied biological tissues - such as human^8^ or animal^9^ dermis, intestinal mucosa^10^, fetal bovine^11^ and bovine pericardium^12^ - each one with relatively different characteristics in terms of clinical and therapeutic applications, described in thousands of scientific publications, in general establishing new standard indications for abdominal wall defects repair, as well as for several other areas.

In Brazil, mainly for economic reasons, there is no literature on acellular matrices for abdominal reconstructions, an unacceptable gap given the high volume of this type of surgery, with evident loss for many patients. Using acellular matrices produced by the company Braile Biomédica, a pioneer in the production of bovine pericardium heart valves in use for 40 years in Brazil, this protocol represents the first national scientific initiative with the use of acellular biological membranes in the abdominal wall.

The main objective of the study was to observe the effectiveness of Bovine Pericardium Acellular Membranes (PeriWall^®^) in the repair of the abdominal wall, analysing the aspects of biocompatibility, tissue neoformation and mechanical resistance through clinical, radiological and histological evaluations. Secondly, compare the results with international publications on the subject, as well as with conventional methods of non-biological synthetic implants.

### Biological Acellular Matrices – Basic Concepts

In the 1990s, the method of deepithelization and decellularization of fresh human skin was developed, with preservation of the basal membrane complex and the structure of the dermis extracellular matrix, thus originating acellularized biological tissues, in this case called acellular dermal matrix (ADM)^13^. Experimental studies showed that the dermal matrix supported fibroblastic infiltration, neovascularization and migration of keratinocytes from a skin graft, also revealing a critical aspect due to the absence of inflammatory cell infiltrate or cell-mediated immune response^14^. These findings quickly led to the use of ADMs in the treatment of burns ^15,16^, representing the first publications with the use of acellular matrices. The observation of its biocompatibility and potential as scaffolds or three-dimensional support for tissue growth cultures, opened the doors to its wide use in bioengineering, evolving to be implanted internally in tissues^17^.

Thus, regardless of their allogeneic or xenogeneic origin, these biological scaffolds are manufactured basically by removing the cellular content of the original tissue, but preserving the three-dimensional structural and functional molecular units of the extracellular matrix (ECM). With different steps and methods adopted by different manufacturers, these ECM extraction methodologies can affect the architecture and biological properties of the generated membrane and may be a factor to interfere in its interaction with the recipient tissue^18^.

An ideal acellular membrane should be composed of collagen and extracellular matrix and not be recognized as a foreign body, due to the absence of a cellular component and absence of all epitopes associated with cells or surface antigens. It would also ideally be recognized as the host’s own tissue in order to induce a minimal inflammatory immune response, allowing it to be revascularized, repopulated by cells and finally incorporated into the recipient tissue^19^. Facing an implanted membrane, three main responses can be observed: integration, degradation / resorption and foreign body reactions. Integration involves the absence of immunological recognition, rapid neovascularization and incorporation, being the ideal response in the presence of biomaterials. The reabsorption is in fact fundamentally the result of enzymatic degradation of ECM by the action of metalloproteases - especially collagenase - consequence of an inflammatory reaction in varying degrees at the membrane / receptor interface, with breakage and progressive elimination of the implant at varying times ^20,21^. The biological scaffolds must withstand a complex balance of resistance to enzymatic degradation while promoting tissue remodeling, gradually degrading until everything that is left behind corresponds to a new and vascularized receptor tissue^22^. Ultimately, no foreign material remains in the long term, minimizing the possibility of chronic inflammatory processes. Foreign body type reactions can occur in membranes that are not fully biodegraded, resulting from the loss of the biological and structural integrity of the MEC in the manufacturing process, causing an immune response that prevents tissue remodelling and integration ^23^.

### Acellular matrices in abdominal wall repair

All biological matrices allow receptor tissue growth in its three-dimensional structure, an important advantage in relation to synthetic implants, particularly in the presence of contamination or infection, where the recurrence rates increase exponentially in abdominal wall repair, by destruction of the implant-fascia interface or even by mandatory explanation for treatment ^24,25^. Another complicating situation refers to the contact of synthetic meshes with the viscera, where the chronic inflammatory response induces visceral tissue growth in the interstices of the implant, often evolving with adhesions, intestinal obstruction or formation of entero-cutaneous fistulas ^26^. These topics have represented the major driver in the increasingly common adoption of biological products, with several studies showing that they have less potential for adhesion to the bowel and greater resistance to infection ^7,27,28^.

Comparative studies between acellular membranes from human and animal origin in abdominal repairs have demonstrated the superiority of xenografts due to their viscoelastic properties and higher resistance matrices. Clinically, abdominal reconstructions with human ADMs show higher rates of protrusion and recurrence of hernias when compared to reconstructions with synthetic mesh, due to the lower resistance to tension and enlargement associated with the increased elastin content in the human dermis^29^. Thus, acellular matrices from porcine, bovine or equine origin were introduced and have been used preferentially in abdominal reconstructions, with forms of cross-linking or not, replacing synthetic meshes in the specific situations described.

In general, publications suggest some superiority of matrices from animal source over human dermal matrices in complex abdominal reconstructions, but there are not enough comparative studies to provide guidelines to guide the choices between the different types. In the vast literature researched here, in only two publications, with level 3 of evidence, the authors report a certain comparative superiority of bovine pericardium over porcine derivatives. Greater intraoperative failure of the bioprosthesis and higher rates of complications in general in the compared application of porcine dermis have been reported, with the indication that the pericardium should be used in reconstructions requiring greater tension during closure^13^.

Discussions regarding the best indications for different pathologies are far from a consensus, but the evolution represented in the field of acellular matrices can be observed by the enormous amount of products available today in the world, with vast literature published, as summarized in Table 1. Not only in the abdominal wall reconstruction, but also in numerous pathologies, bioprosthesis seem to represent a new era in the application of biomaterials and should be incorporated into the therapeutic armamentarium of surgeons.

**Table 1.**
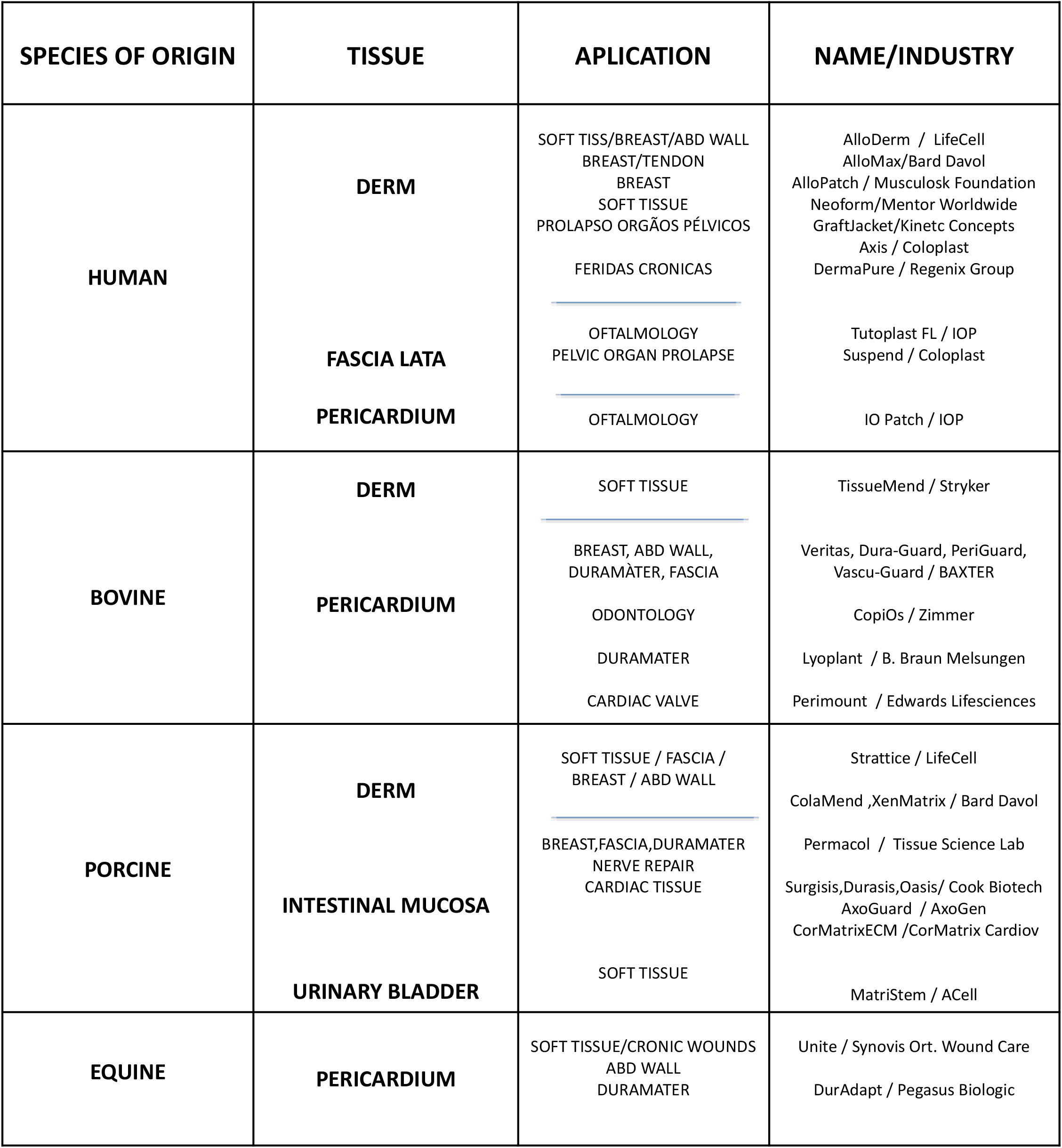
Partial list of the main acellular matrices from human and animal origin. All of these products are approved and available around the world market for their different applications, with hundreds of scientific publications, demonstrating the evolution and growing importance of the therapeutic application of bioprostheses.

## MATERIALS AND METHODS

Thirty patients underwent abdominal wall repair associated with bovine pericardial membrane (PeriWall^®^), 14 men and 16 women, aged between 29 and 77 years (mean = 48 years), for different indications and locations, making a total of 40 anatomically individualized implants, summarized in table 2. The minimum follow-up was three months and the maximum 38 months, with an average of 22 months.

**Table 2.** Repaired abdominal wall deformities with bovine pericardium acellular matrices, with a total of 30 patients and 40 anatomically individualized implants.

| **PATHOLOGY** | **CASES (N)** | **IMPLANTS (N)** |
| --- | --- | --- |
| **HERNIAS** |  |  |
| **INGUINAL UNILATERAL** | 10 | 10 |
| **INGUINAL BILATERAL** | 03 | 06 |
| **INCISIONAL** | 04 | 04 |
| **UMBILICAL/PARAUMBILICAL** | 02 | 02 |
| **EPIGASTRIC** | 01 | 01 |
| **POS BARIATRIC/ ABDOMINAL WALL BULGING** | 06 | 09 |
| **ABD WALL ENDOMETRIOSIS - INFRA UMBILICAL** | 01 | 01 |
| **ASSOCIATED DEFORMITIES** |  |  |
| **INGUINAL + SUPRA UMBILICAL HERNIA** | 01 | 02 |
| **POS BARIATRIC + SUPRA UMBILICAL HERNIA** | 01 | 02 |
| **ABDOMINAL WALL BULGING + BILATERAL ING HERNIA** | 01 | 03 |
| **TOTAL** | **30** | **40** |

## METHOD

In a multicentre character and approved by the Research Ethics Committees, patients were selected and operated on by the main author and the general surgery teams with expertise in the area in three reference hospitals in São José do Rio Preto(São Paulo,Brazil) namely: Hospital do Coração do Instituto de Moléstias Cardiovasculares (HC-IMC), Associação Portuguesa de Beneficência and João Paulo II State Hospital. All patients received informed consent about the nature of the procedure, describing the mandatory need for an implant mesh to correct the problem and the options between the synthetic and the biological compound.

In addition to the main pathology and classification of the surgical wounds nature^32^, the variables age, Body Mass Index and possible associated pathologies were recorded, important for the inclusion / exclusion criteria. The only inclusion criterion corresponded to the standard indications for synthetic non-biological meshes use, that is, in surgeries in which the implantation was mandatory. Preoperatively were excluded obese patients, patients with contaminated or infected surgical wounds and patients with important associated comorbidities (obesity, arterial hypertension, diabetes, emphysema and / or obstructive pulmonary disease), with a risk-benefit ratio considered unfavourable by the medical team.

All patients underwent surgery under general anaesthesia, with a mean hospital stay of 1 day, and received antibiotic therapy with cephalosporin 1g per day for 7 days. The use of compressive garments for a period of 120 days was recommended, as well as a physical exercise during this period.

In addition to monthly follow-up, radiological exams were performed by ultrasound at different postoperative periods and, in 04 cases, Electromagnetic Resonance of the abdominal wall at 09, 11, 17 and 26 months.

In 03 patients it was possible to perform biopsies of the implanted areas at 13, 22 and 23 months post op and the material stained in Hematoxylin-eosin, Gomori’s Trichrome and Picrosirius red.

### Implants preparation

The membranes were supplied sterile in flasks containing 4% formaldehyde requiring pre-implantation washing, as recommended by the manufacturer. The membrane was washed with 0.9% saline, discarding the solution every 5 minutes and repeating the procedure for 3 cycles, requiring 15 minutes for the material to be ready for use. The principles of “no touch” were followed when handling implants, changing and washing gloves to remove talcum residues and new surgical site asepsis was made prior to implantation. A solution was prepared containing 1,000 ml of 0.9% saline associated with 2 g of cephalosporin and 80 mg of gentamicin, leaving the washed membrane immersed in this solution until the moment of its use. During the fixation process, the same solution was used to irrigate the entire implantation area at random times, keeping the surgical field as clean as possible.

### Surgical Technique

Different techniques were used depending on the complexity and location of the abdominal reconstruction, also varying according to personal choices, but following standard methods in the preparation and manipulation as well as at the implant’s fixation. The membranes were fixed with separate and / or continuous stitches of non-absorbable polypropylene (Prolene^®^) 2-0 or 3-0 threads on their periphery, complemented with mattress sutures on their surface. Routinely, absorbable stitches of Polyglycolic Acid 2/0 (Vicryl^®^) or Polydioxanone (PDS^®^) 2/0 were applied from the subcutaneous to the membranes and muscle aponeurosis over the entire length of the detached area, immobilizing the flap and achieving maximum reduction of dead spaces, preventing seromas and promoting the largest possible contact surface at the membrane-tissue interface **(FIGURE 1**). In all fixations, care was taken to avoid exaggerated folds in the implant, being stretched with maximum tension.

**FIGURE 1.**
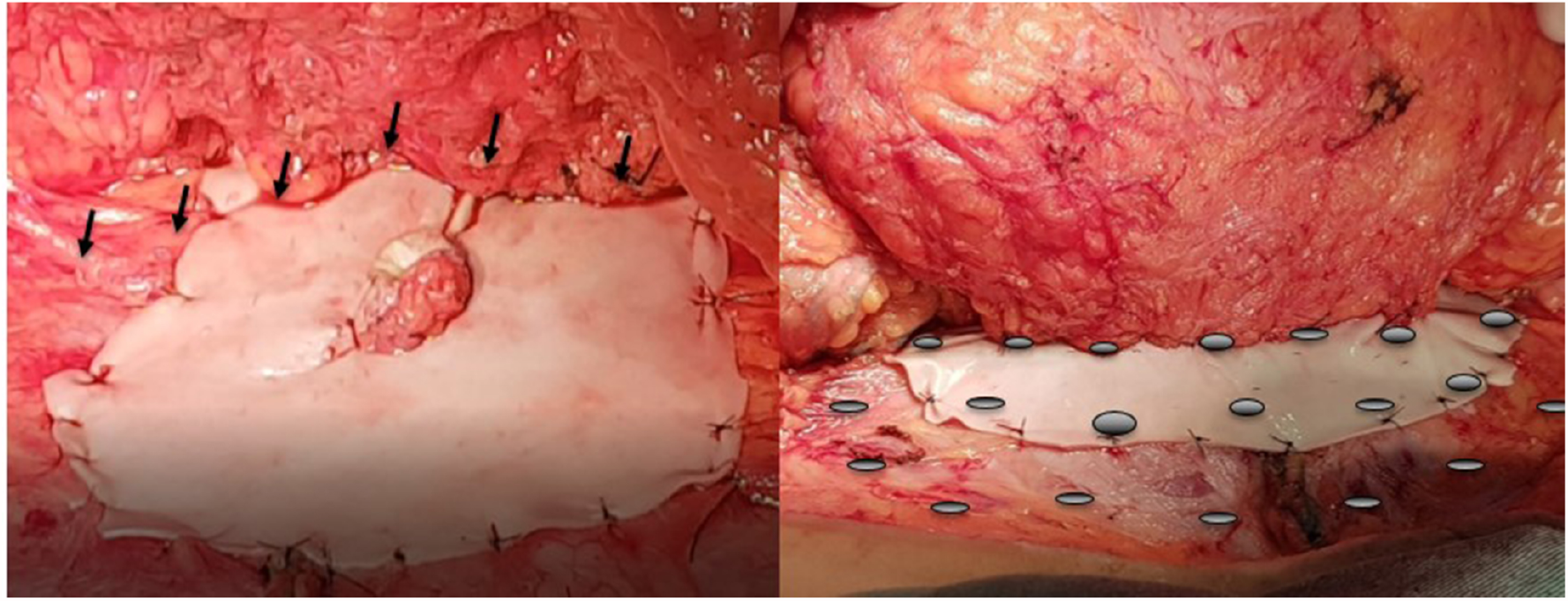
**Intraoperative** view of a suprafascial membrane fixation to the subcutaneous of the abdominal wall. **Left:** Vicryl 2/0 separate stitches are passed across the entire length of the detachment, fixing the subcutaneous to the membrane and abdominal wall every 2 to 3 cm (black arrows), repeating the procedure inferiorly every 2 to 3 cm until the final suture border (gray circles on the **Right**). This maneuver allows a total immobilization of the flap with maximum dead spaces reduction, promoting a greater contact surface of the membrane-tissue interface and preventing seromas. In none of the operated cases, aspirating drains were used.

In inguinal hernias (19 cases), the membranes were fixed to the inguinal canal with separate stitches of Prolene 2/0 and kept buried in a subfascial position. (**FIGURE 2A**). When closing the aponeurosis, separate points of Vicryl^®^ 2/0 were given anchoring the membrane as described, maintaining the final closure without using drains. The average size of the implants was 10 x 6 cm.

**FIGURE 2.**
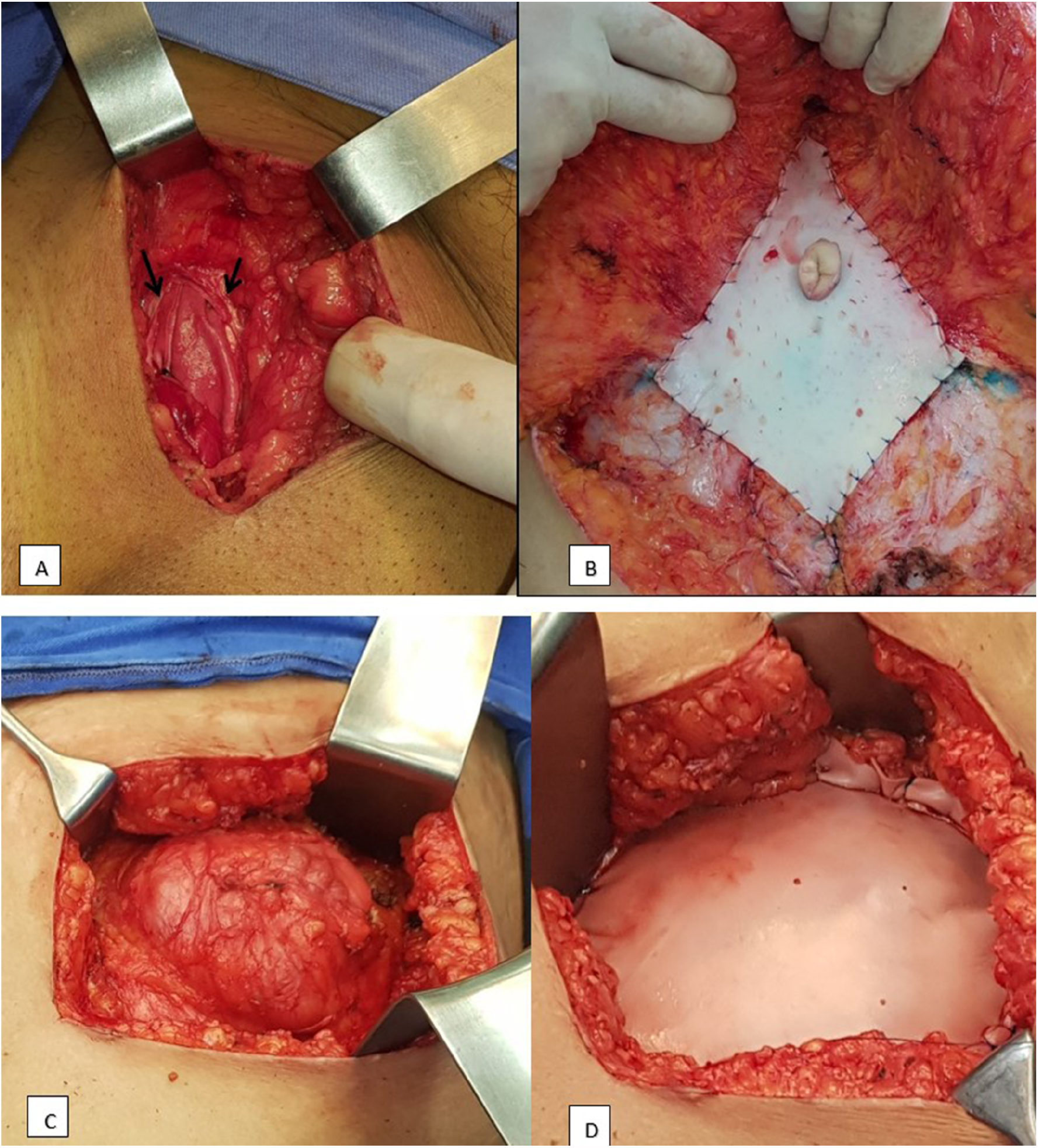
**A-** Inguinal hernia repair on the left. View of the membrane fixed in the inguinal canal, below the muscular fascia (black arrows) that will be sutured, with the implant in a subfascial position. **B** - Implant of acellular membrane in suprafascial position, directly sutured over the muscular fascia after primary muscle plication. **C** - Intraoperative aspect of an incisional hernia with a large defect (about 10 cm in diameter), impossible a direct closure of the muscular layers. **D** - Correction of the defect with bridged implant, fixing the membrane under tension externally at the edges of the defect, directly covering the peritoneum.

In umbilical and epigastric hernias (05 cases) it was possible to make the direct primary closure of the musculature and aponeurosis, with separate points of Poliglecaprone 1/0 (Caprofyl^®^) and the membrane being affixed in a suprafascial position, sutured covering the area treated with separate stitches of Prolene 3/0 **(FIGURE 2 B**). After the membrane fixation routine, the final closure was maintained without the use of drains. The average size of the implants was 6×6 cm.

In incisional hernias, the membranes were also sutured in a suprafascial position, except in 01 case where the primary approach of the musculature was not possible (**FIGURE 2 C, D**). In these cases, the membrane used had an average dimension of 10×10cm.

In repairs due to exaggerated bulging of the abdominal wall (08 cases) and immediate reconstruction after tumour resection (01 case), after muscle plication with absorbable threads of Caprofyl^®^ 1-0 or polydioxanone (PDS^®^), the membranes were sutured in suprafascial position with separate stitches of Prolene 3/0. These cases required the largest membranes - 15×10 cm - as there was also a need to use more than one membrane, with supra and infra umbilical positioning in 3 cases. The fixation of the subcutaneous tissue to the surface of the membranes was also performed, eliminating the use of aspiration drains in all cases. (see **Supplemental video 1** demonstrating the described procedures).

## RESULTS

The patients had good results, with no deformities recurrence, either clinically or radiologically. There was also no record of hematomas, infection or any phenomenon of a local or systemic reaction nature. In one post-bariatric case, there was an infra-umbilical seroma, treated by aspiration puncture with removal of 60 ml of secretion without other recurrences.

Two patients had scar revision at 13 and 22 months postoperatively and 01 muscular wall revision post endometrioma resection at 23 months. In all of them it was not possible to make any visual identification of the implanted membranes, with the surgical area of normal appearance, without reaction areas and with mild surgical fibrosis. In all of them, biopsies were made at the implantation area **(FIGURE 3**).

**FIGURE 3.**
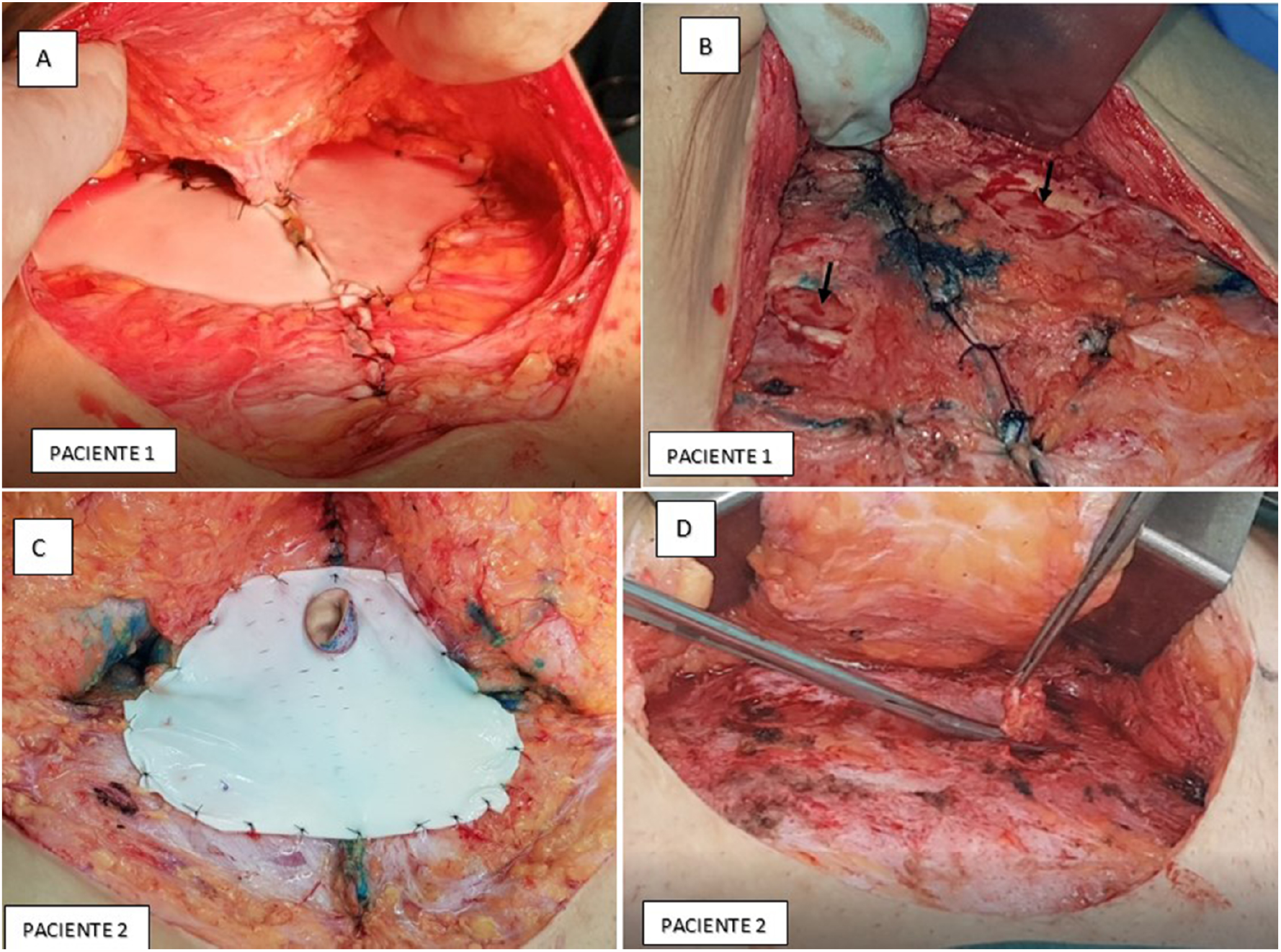
On the **left**, in **A** and **C**, final aspect of abdominal wall repairs associated with acellular bovine pericardium, sutured in a suprafascial position after muscle plication. On the **right**, the intraoperative aspect of surgical revision in the same previously repaired site, in **B** in patient 1 at 22 months and in **D** in patient 2 at 23 months postoperatively. In both reviews there was a normal cicatricial aspect at the previously implanted area, with the absence of foreign body reaction or granulomas, with no visual evidence of residual membrane. Tissue samples for histological analysis were taken in the areas corresponding to the location of the previous implant (black arrows in B and removing the sample with scissors in D).

### Radiological assessments

In all analysed periods the Ultrasonography did not allow any identification of the implanted membranes. In the first 30 postoperative days, small sparse seromas can be seen in the area of the implants, without clinical relevance, disappearing after this period. There were no long-term local recurrences, as well as anatomical changes in the operated region (FIGURE 4).

**FIGURE 4.**
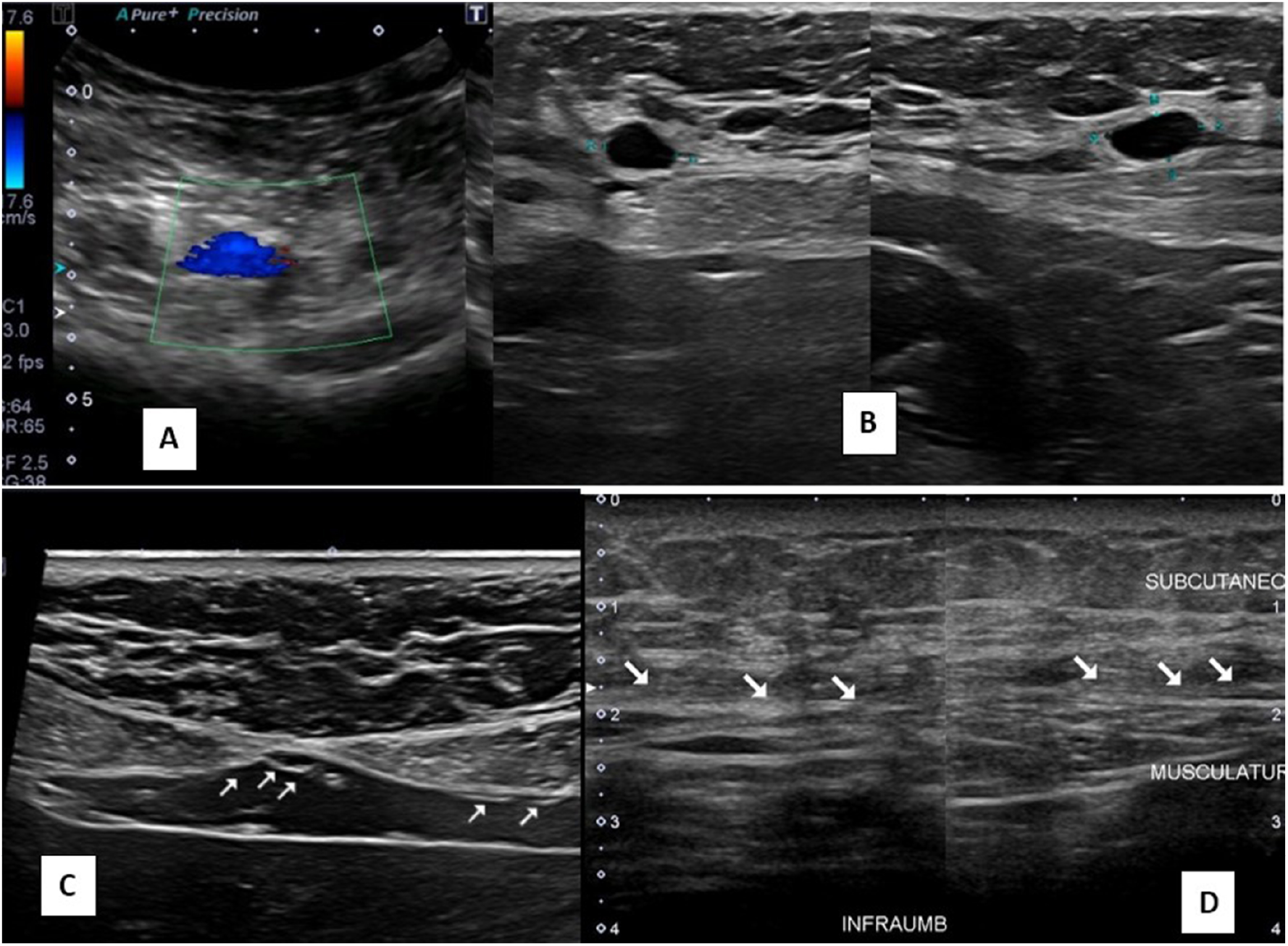
Abdominal wall Ultrasound images, post repair with acellular pericardium matrix, in different postoperative periods. Images in **A**, at 15 days after inguinal herniorrhaphy and **B**, at 30 days after incisional herniorrhaphy, showing small isolated seromas, without clinical repercussion or need for drainage. In **C** and **D**, images at 45 and 150 days showing normal appearance after incisional hernias repair, with no recurrence or anatomical changes. The arrows identify the subcutaneous interface / muscle aponeurosis, with a usual anatomical aspect. In any case the technique allows visualization of the implanted membranes.

In the electromagnetic resonances, performed at 09, 11, 17 and 26 months after the operation, it was also not possible to identify the membranes in the implantation site or indirect signs of their presence. No local recurrences or major anatomical changes in the operated areas were identified. (FIGURE 5)

**FIGURE 5.**
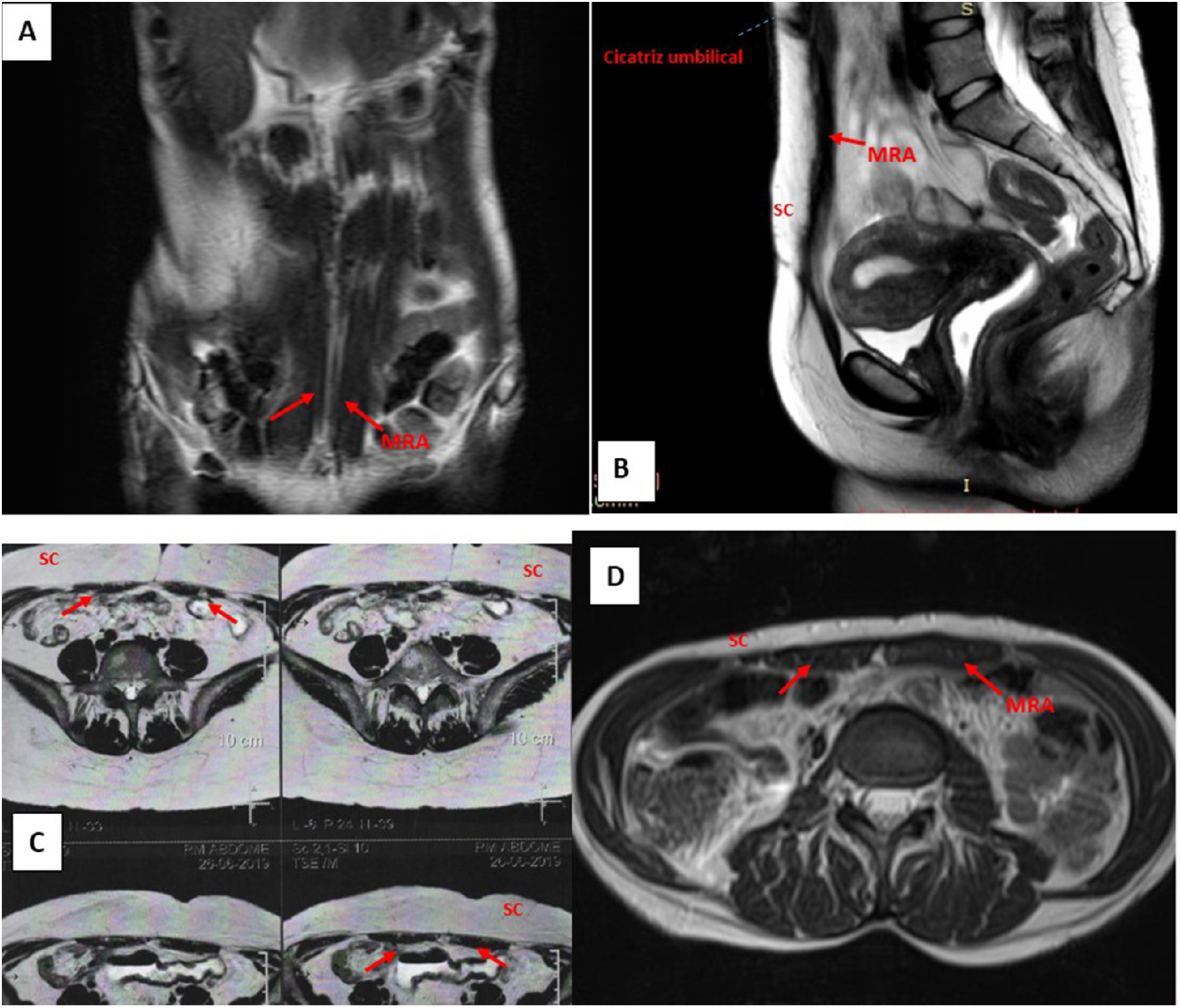
Abdominal wall Electromagnetic Resonance images, post repair with acellular pericardium matrix, in different postoperative periods. In **A**, 9 months post-bariatric abdominoplasty; in **B**, 11 months after repair by resection of infra-umbilical wall endometrioma; in **C and D**, at 17 and 26 months after incisional hernia repair. In all cases the membranes were fixed in a suprafascial position and the slices correspond to the implantation areas. In **A, C** and **D**, normal abdominal wall anatomy is observed, with absence of diastasis of the rectus abdominis muscles (red arrows), seromas, herniations or other changes. In **B, C and D**, the subcutaneous / aponeurosis interface is clearly observed, presenting a normal aspect, and it is not possible to identify the presence of the implanted membranes. (SC - SUBCUTANEOUS; RAM - RECTUS ABDOMINIS MUSCLE)

### Histological assessments

All samples showed important tissue neoformation replacing the implanted membranes, with important deposition of collagen and normal-looking cellular tissue. No important local reactions were observed, with some isolated focal points showing macrophages in a mild inflammatory process. In all periods analysed it was still possible to identify, in a small amount, the presence of fragments of acellular tissue corresponding to the original membrane (FIGURE 6).

**FIGURE 6.**
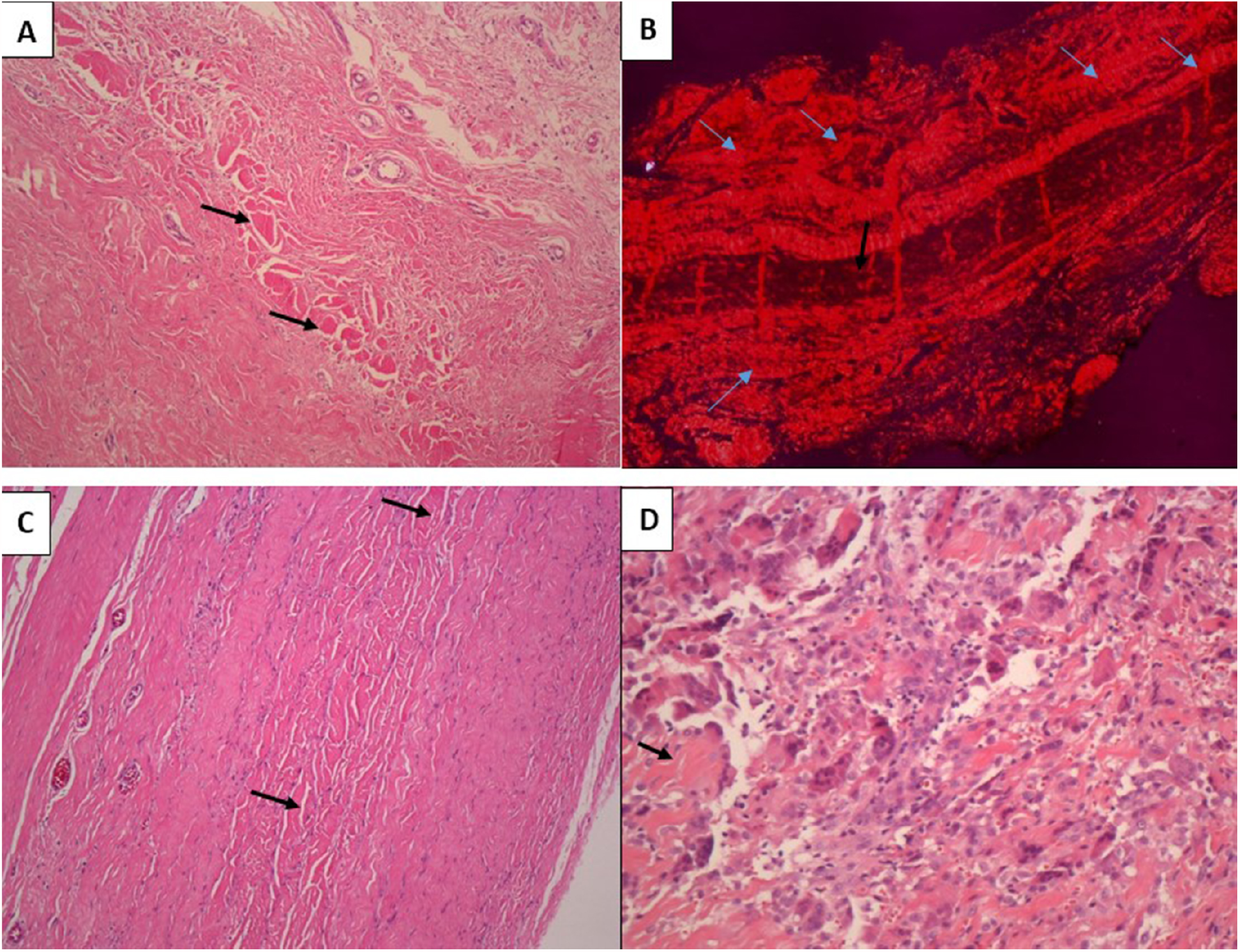
Histological sections from areas corresponding to suprafascial abdominal implants, in the postoperative periods of: **A and B**, 13 months; **C**, 22 months and **D**, 23 months. In all periods normal-looking cellularized tissue is observed, replacing the implanted matrices, without significant inflammatory activity or foreign body-type reactive granulomas. In **B**, there is a marked deposition of collagen in the degraded areas of the extracellular matrix (blue arrows). In all samples, fragments of decellularized tissue from the implanted membrane are still observed (black arrows), in greater quantity at 13 months postoperatively. Hematoxylin-Eosin staining in **A, C and D**. Picrosirius Red staining in **B**. Increase 40xx in A, B and C; Increase 100xx in D.

## DISCUSSION

Biological scaffolds have been increasingly used not only in the abdominal wall reconstruction but also in several therapeutic applications, revealing a conceptual evolution in biomaterials application. The nature of the three-dimensional molecular organization distinguishes extracellular matrix scaffolds from synthetic material due to the possibility of repair by tissue remodelling instead of scar fibrosis, strategies pursued by the concepts of tissue engineering and regenerative medicine^33^.

Although there is still no consensus as to which bioprosthesis would be ideal for abdominal wall repair, bovine pericardium has been successfully adopted for this purpose since it has advantages over synthetic materials, due to its biocompatibility and physical characteristics of resistance and malleability. It has been shown that bioprostheses have an increased ability to integrate with surrounding tissues, demonstrating resistance to infection, extrusion, erosion and adhesion formation^34^. Thus, acellular membranes have been particularly indicated to replace synthetic implants facing more complex cases, in the presence of contaminated wounds and in situations of direct contact between the implant and the viscera^35^.

The presented outcomes suggest that the employed bioprosthesis showed a behaviour similar to that documented in the literature, showing efficacy in the correction of the defects with no record of recurrences or adverse events, in addition to presenting excellent biocompatibility and integration with adjacent tissues. It was possible to directly observe the total incorporation of the implants at the revisions, in addition to the described results in radiological and anatomopathological evaluations.

In the adopted protocol it was not possible to evaluate the matrices in more complex repairs and in contaminated wounds, but it can be assumed that, due to the similarity presented by the material in the numerous available publications, its use can be considered without restrictions in future indications in abdominal wall repairs. The proposed protocol deliberately limited the indications to cases of lower risk, both from the point of view of the pathology and patient clinical conditions, classified as Grade I^36^, limiting factors of comorbidities that could interfere in the analysis of the implant itself.

In addition to the deformity recurrence, events classified as “occurrences in the surgical site” - especially infections and seromas - are the main problems after corrections of ventral hernias, with reports estimating that up to 75% of recurrences are due to infection and inadequate fixation of the implant^37^, indicating prevention as paramount. The methodology adopted was totally focused in this sense - prevention of infections, seromas and adequate fixation of the implants - and should be an important factor corroborating for the absence of adverse events in this series.

Regarding infection, the special care described in handling the implants and at the implantation site were followed as a permanent routine, following the successful routine adopted many years ago in breast implants, summarized under the concept of “no touch” in contamination and biofilm formation prevention^38^. Conceptually a biological event diverse from infection^39^, eventual microbial biofilms formation on the surface of abdominal meshes - of a synthetic or biological nature - is a factor to be considered in reactive events in the postoperative period^40^, and should be considered an etiological agent to be fought with strict protocols in the intraoperative.

The same applies to seroma prevention and rigorous implants fixation, a routine used in any type of biomaterial selected. The progressive sutures of the subcutaneous layer of the abdominal flap to the implant and muscle fascia surface is a smart strategy fulfilling several objectives in an efficient way, namely: 1.promotes flap and implant immobilization, preventing displacements while amplifying the membrane / tissue interface contact, favouring the biological processes of wound healing; 2.reduces dead space preventing seroma and simultaneously eliminating the use of drains. Aspiration drains were not used in any of the cases in this series and in only one case there was a need for aspiration puncture to treat a small seroma, solved in a single session. Postoperative ultrasonographies show small sparse seromas present in the first 30 days, not being observed in later cases and showing a normal-looking subcutaneous / aponeurosis interface, without visualizing the membranes. In addition to being more comfortable for the patient and the nursing staff for dressings, the use of drains can also favour contamination of the surgical site and its effectiveness in preventing seromas is not properly demonstrated^41,42^. The results obtained in this series demonstrate that the strict fixation of the subcutaneous / aponeurosis / implant interface, with progressive sutures of the detached area, does not require the use of aspiration drains in abdominal wall repairs.

Technical aspects of the implants positioning in relation to the muscular plane - subfascial, suprafascial, submuscular or bridged - are described factors that can also increase the risk of complications ^43,44^. In this series, the membranes were implanted in the subfascial space for inguinal hernias and suprafascial with primary muscular closure in incisional hernias and post bariatric protrusions, positioned in bridge in only 01 case. Aided by the fixation methods, the results show that suprafascial positioning of bioprostheses is a simple and effective method for the correction of less complex hernias, as described in other publications^45^.

Biological scaffolds must be degraded by action of metalloproteases^46^ and replaced by native tissues over time, serving only as a temporary structure for host cell growth, a process defined as remodelling. The clinical utility of these materials depends on the balance between the degradation rate and internal growth of native tissue. If the implant is absorbed before adequate neovascularization, tissue growth and differentiation / collagen deposition processes, the overall quality and strength of the newly formed tissue are likely to be insufficient to repair the abdominal wall. In this context, the discussions between the nature of cross-linked and non-cross-linked materials are focused, with different properties with regard to incorporation, resorption and mechanical resistance processes, a still controversial topic in abdominal wall repair ^47,48^. Publications describe the non-cross linked materials as being of faster incorporation and resorption^49^, with the crosslinking prolonging the degradation time and conferring greater degrees of resistance to traction and stretching, in addition to resistance to infections ^50,51^, factors that directly interfere in the healing aspects at the implant site. Experimental studies have shown that the crosslinking with glutaraldehyde conferred some resistance to the activity of collagenase ^52^, with several bacteria also showing important collagenolytic activity^20^. This could justify the claim that cross-linked bioprostheses must be relatively more resistant to bacterial degradation and, therefore, safer for use in contaminated environments^53^. Studies with decellularized bovine pericardium, submitted to different degrees of crosslinking, showed a direct relationship between resistance to enzymatic degradation and crosslinking degree, demonstrating that this aspect determines the rate of degradation of the acellular tissue and its pattern of tissue regeneration ^54,55^. Due to these factors, it is believed that crosslinking increases the durability of implanted biomaterials, conferring a greater capacity to provide adequate support for remodelling processes with endogenous collagen in abdominal hernias repair ^48^.

The acellular pericardium matrix employed in this study, with crosslinking by glutaraldehyde, showed similar properties to other bioprostheses reported in terms of biocompatibility and progressive degradation of the material. In revisional cases, adhesions or important inflammatory processes were not observed in the surgical sites, and it was not possible to visualize the previously implanted membranes, fully incorporated. Histological analyses showed normal aspects of remodelling with normal tissue growth and collagen deposition, with progressive degradation of the implanted membranes, present in greater quantity at 13 months and still residual in samples of 22 and 23 months.

The choice between using a synthetic versus biological material for a given application depends on several factors, including the procedure cost-benefit relation. At a much higher cost, acellular membranes are currently restricted to more complex cases, but the financial aspect cannot limit the proper understanding of the conceptual evolution underlying the study and use of biological materials. Extracellular matrix membranes should be seen as a microenvironmental niche with structural and functional components preservation and retained biological activities in tissue regeneration processes ^56^, going beyond the issue of a structural scaffold or just a mechanical support device. With the recognition of the presence of the implant, the processes of cell degradation by the recipient begin with consequent release and recruitment of signalling molecules as growth factors, nanovesicles linked to the matrix, cytokines and chemokines^57^. This process has an important impact on local biological activity, with recruitment of stem and progenitor cells^58^ and the modulation of the innate immune response^59^, favouring an environment of structural and functional remodelling instead of the formation of a fibrous capsule of scar tissue or chronic inflammatory processes.

Thus, unlike the exclusively mechanical role of synthetic implants, bioprosthesis play an active role in biological events at the implantation site, pointing in the direction of regenerative processes that must be properly known and experienced by surgeons, whose experience is estimated as one of the most important prognostic factors in the repair of abdominal wall hernias ^60^.

## CONCLUSIONS

The acellular pericardium matrix analysed showed efficacy, with no recurrences or adverse events, presenting biocompatibility and integration with adjacent tissues. It was possible to observe in situ the implants incorporation, in addition to microscopically demonstrating their progressive biodegradation, with collagen deposition and tissue neoformation in the recipient bed. Due to the similarity presented to other matrices described in literature, the material can also be considered in complex abdominal wall reconstructions, as well as in other possible applications.

The adoption of strict protocols in implant manipulation and fixation can positively impact the results of abdominal wall repairs. The progressive sutures of the subcutaneous / implant / aponeurosis interface should be a routine when handling implants of any nature, preventing biofilms formation and the use of aspirative drains.

Biological scaffolds bring important conceptual evolutions that must be incorporated by surgeons, with bioprosthesis choice being considered by factors that go beyond the procedures’ cost-benefit. Series with a greater number of cases and in complex reconstructions will better define the criteria for their indication.

## Supporting information

DISCLOSURE FORM

SUPPLEMENTAL VIDEO 1https://youtu.be/lRWVo_snDuY

## Data Availability

all data contained in this manuscript are available for requests and clarifications at
SUPLEMENTAL VIDEO1: https://youtu.be/lRWVo_snDuY

